# What pushed Israel out of herd immunity? Modeling COVID-19 spread of Delta and Waning immunity

**DOI:** 10.1101/2021.09.12.21263451

**Authors:** Hilla De-Leon, Dvir Aran

## Abstract

Following a successful vaccination campaign at the beginning of 2021 in Israel, where approximately 60% of the population were vaccinated with an mRNA BNT162b2 vaccine, it seemed that Israel had crossed the herd immunity threshold (HIT). Nonetheless, Israel has seen a steady rise in COVID-19 morbidity since June 2021, reaching over 1,000 cases per million by August. This outbreak is attributed to several events that came together: the temporal decline of the vaccine’s effectiveness (VE); lower effectiveness of the vaccine against the current Delta (B.1.617.2) variant; highly infectiousness of Delta; and temporary halt of mandated NPIs (non-pharmaceutical interventions) or any combination of the above. Using a novel spatial-dynamic model and recent aggregate data from Israel, we examine the extent of the impact of the Delta variant on morbidity and whether it can solely explain the outbreak. We conclude that both Delta infectiousness and waning immunity could have been able to push Israel below the HIT independently, and thus, to mitigate the outbreak effective NPIs are required. Our analysis cautions countries that once vaccines’ will wane a highly infectious spread is expected, and therefore, the expected decline in the vaccine’s effectiveness in those countries should be accompanied by another vaccination campaign and effective NPIs.

## INTRODUCTION

First discovered in India in October 2020, the Delta variant became the dominant variant by March 2021 [1], and caused the deadliest surge to date, with over 200,000 documented COVID-19 associated deaths in two months [2]. Delta quickly became the dominant variant in most countries after introduction, replacing all other variants of concern (VOC) [1]. The rapid dominance of Delta is attributed to its high infectiousness, and is predicted to have a basic reproduction number, *R*_0_, of 3.2 to 8, with a mean of 5.08, as opposed to Alpha, which was estimated to be approximately 4 [3]. A recent study confirmed that the viral load of individuals infected with Delta is significantly higher than Alpha [4].

Vaccination roll-out in Israel of the COVID-19 vaccines started on December 20, 2020. By the end of February 2021, over a third of the whole population and over 80% of adults over the age of sixty, were already fully vaccinated by the BNT162b2 vaccine developed by BioNTech and Pfizer [5]. This successfully conducted vaccination campaign resulted in a drop in cases in Israel from over 1,000 per million per day to less than 3 cases per million per day in May 2021. By May, Israel was thought to have reached the herd immunity threshold (HIT), which prompted the lifting of some non-pharmaceutical interventions (NPIs), including a decision to halt mandated masking by mid-June.

Nevertheless, a new outbreak began in mid-June, first, with dozens of infections coming from abroad, triggering some outbreaks within local communities that went on to spread across the country. By the end of August, Israel went back to over 1,000 confirmed cases per million per day. This outbreak, from its very beginning, was completely dominated by infections with the Delta variant. Additionally, since the beginning of the outbreak, a high increase in breakthrough infections in vaccinated individuals was reported. Later, the waning of immunity was confirmed in multiple studies worldwide, showing that the BNT162b2 vaccine effectiveness (VE) is reduced to below 50% after five months of the second dose [6–9]. Note that decline in VE is most prominently observed for infection, and not for hospitalization, severe disease and death, where only mild decline is observed. Finally, the Israeli government reinstated some NPIs in late June, including indoor masking, green passports and mandatory isolations.

What caused this surge to start in Israel in June 2021 or what propelled Israel below the HIT is a hot topic of debate. In the HIT formula, both the *R*_0_ and the fraction of protected individuals are taken into account [10]. Consequently, both the higher infectiousness of Delta and the reduced effectiveness of the vaccine in those vaccinated in January and February may have pushed Israel below the HIT. Using a novel spatial-dynamic model of disease spread, which has shown high success in modeling the spread in Israel throughout the COVID-19 pandemic [11, 12], we compared models of infections that combine an *R*_0_ of Alpha or Delta and decline and no decline in the effectiveness. Our analysis suggests that the current outbreak in Israel is caused by a combination of both the Delta variant being more contagious than the Alpha variant and the decline of VE as time passes. However, we find that even in the presence of the Alpha variant, the decline in VE would have pushed Israel below the HIT. Moreover, we found that the Delta variant itself would push Israel below the HIT even without waning immunity, but with a different age mixture of confirmed cases compared to the scenario in which there is waning in the effectiveness but the dominant variant is Alpha.

## METHODS

In this work, we use a diffusive Monte-Carlo (MC) model introduced by De-Leon and Pederiva [11, 12] to model the spread of the disease. We used this model to examine the expected effects of vaccinations on both confirmed cases (CC) and severe hospitalizations, as well as to estimate the vaccine’s current effectiveness. As opposed to other infection models, such as the Susceptible Infected Removed (SIR) [13–15], this particle model enables us to distinguish between different age groups and treat each one separately, assuming that the infection occurs throughout the population simultaneously. Furthermore, a particle model can be adjusted to adapt the rate of population immunization to accurately examine the different effects of the vaccine on subgroups of the vaccinated population as well as the entire population.

It is necessary to differentiate between *R*_*t*_ (the theoretical replication number of the virus) and *R*_*e*_ (the effective replication number of the virus) in order to estimate the spread of a pandemic in the presence of vaccines. *R*_*t*_ estimates the number of encounters between carriers and healthy individuals that would have resulted in an infection if the vaccines had not been available and is defined by how contagious the current variant is, whereas, *R*_*e*_ is affected by vaccination rates and protection offered by the vaccines. In this work, based on the easing of social restrictions in Israel in February 2021 and Delta’s increasing dominance since June, we estimate the theoretical *R*_*t*_ in Israel in July 2021 at 3 for the Delta variant and 2.2 for the Alpha variant (we perform the modelling on additional *R*_*t*_ levels). It is important to note, that in contrast to *R*_0_, the basic reproduction rate, *R*_*t*_ takes into account the NPIs that are still customary in Israel, especially the duty of isolation for verified patients and those who have been exposed to a verified patient and the duty of wearing masks indoors.

Recent studies from Israel, Qatar, UK and [6, 9, 16–18] have shown reduction in effectiveness in preventing COVID-19 infection and transmission blocking in individuals vaccinated more than 5 months before exposure. While the exact effectiveness after this waning humoral immunity, reports from Israel have estimated the effectiveness of the vaccine against infection to be between 20%-50% for those vaccinated six months ago, compared to 80%-90% for those vaccinated recently.

Data on deaths, vaccination rates and population size of countries was download on September 1, 2021 from OurWorldInData.org [**?**]. Only countries with a population over 1 million and with available information are presented. Data on deaths and population size of USA counties was downloaded on September 19, 2021 from [19]. Data on vaccination rates was downloaded on September 19, 2021 from [20]. Counties with 0 deaths in August or 0% vaccinations on April to July were removed, assuming incomplete county level information (only 40 states had information stratified by county level).

## RESULTS

After a wide vaccination campaign in Israel, the number of new confirmed and hospitalized cases declined from mid-January 2021, first among adults over 60 years of age, and later in the entire population [21, 22]. After almost complete elimination of cases, a new outbreak began in mid-June 2021. We applied our spatial-dynamic MC algorithm with information on current vaccination rates, COVID-19 recoveries, and the number of confirmed cases in Israel from July 2020 to June 2021 to model difference scenarios of the expected daily confirmed cases in the outbreak that started in June 2021 (Figure 1). We first ran our model to estimate cases in the presence of the Delta variant and in time-dependent decline of VE. This model performed well in predicting the real-world counts of CC (normalized mean absolute difference = 2%). We next ran a model with a theoretical reproduction rate (*R*_*t*_) that corresponds to the Alpha variant, and also without a time-dependent decline in VE. This model suggested that if both factors have not happened, Israel would still be below the HIT, and the June outbreak could have been contained by the vaccinations alone. We next tested two additional models: a model with Delta but no time-dependent decline in VE (Delta model), and a model with Alpha and with a time-dependent decline in effectiveness (Waning model). These models show that both factors - more infectious variant and waning immunity - were able to push Israel below the HIT independently. In both models, the number of CC would have grown steadily up to approximately 2000 daily cases in the Waning model and 3,500 daily cases in the Delta model by mid-August. According to our analyses, with decline in VE, an *R*_*t*_ below 2 could have allowed to contain this outbreak without additional NPIs, or at least delay it (Supplementary Figure 1).

**FIG. 1:**
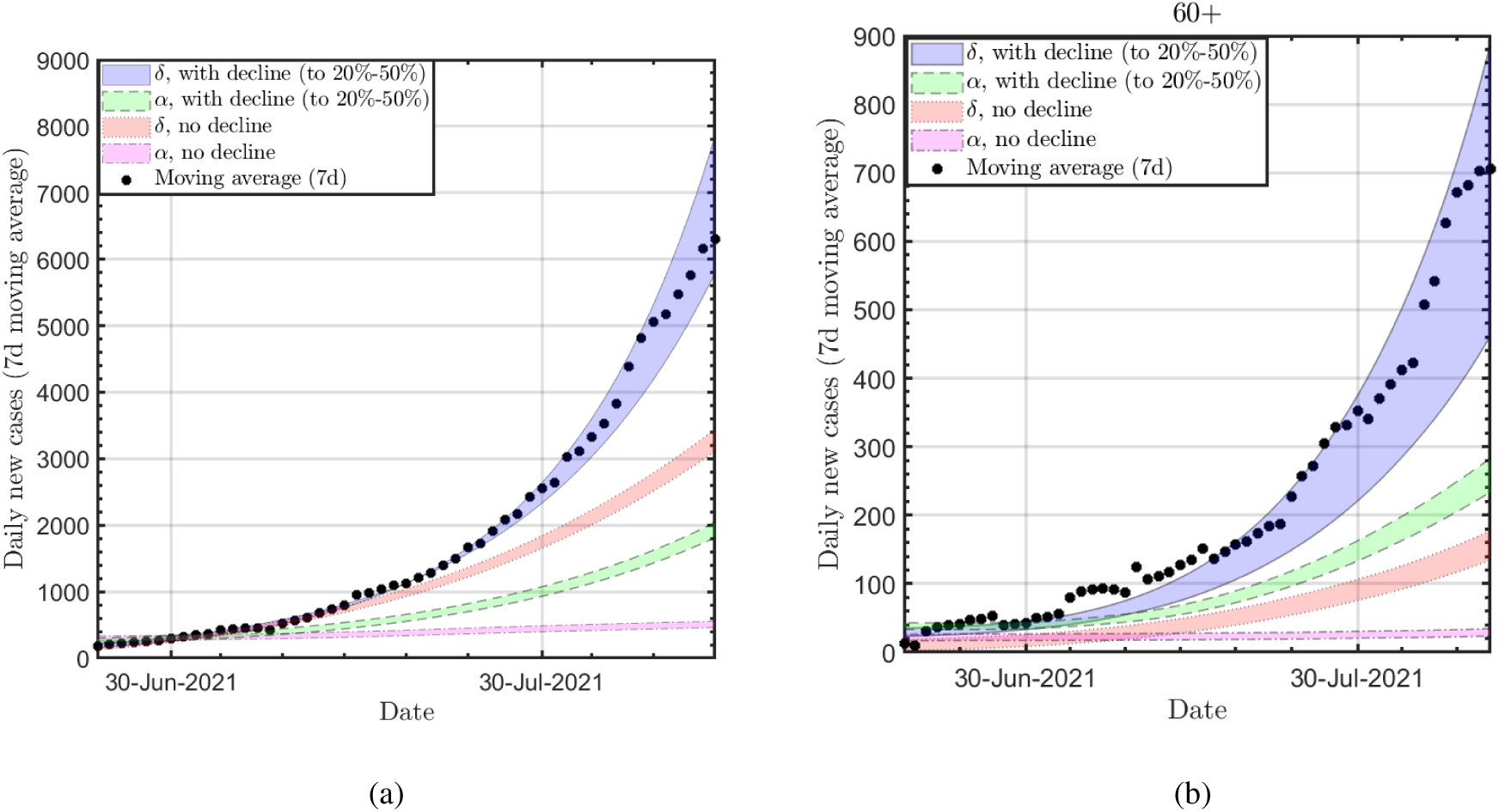
Predicted daily cases in four scenarios. The daily new cases (a, total population, b 60+). Solid blue band: the delta variant with time-decline of the effectiveness against infection to 20%-50% 150 days after the second dose; Dashed green band: alpha variant with time-decline of the VE against infection to 20%-50% 150 days after the second dose; Dotted red band: the delta variant without a time-decline of the VE against infection; Dotted-dashed magenta band: the alpha variant without a time-decline of the VE against infection. Dots: Observed data (7-day moving average).

Interestingly, due to the fact that in the Waning model the reduced VE is expected first in the elderly population who were vaccinated first, the number of CC in 60 year old and over individuals is predicted to be higher in the Waning model compared to the Delta model (250 cases vs. 150 cases in mid-August) (Figure 1b). In the Delta+Waning model, this number grows to 700 cases (in the real-world there were 716 cases). By examining the age distribution of cases at the beginning of the outbreak, we found that our prediction matched what actually occurred. While during the winter outbreak the fraction of cases among adults 60 and over was significantly lower than their fraction in the total population (10% of cases vs. 15% of the total population, One-sample proportional test p-value = 0.009) due to the higher vaccination rates in older populations, in the June outbreak we observed an increased percentage in the adult population that were affected (17% vs 15%, one-sample proportional test p-value = 0.7), more or less as expected from random spread in the population (Supplementary Figure 2).

Since the Delta variant became the dominant variant in most countries by July 2021, and waning immunity was observed in other vaccinations, it is anticipated that countries with low vaccination rates or that have vaccinated their population early, will experience a spike in infections in August. On the other hand, countries that have only recently vaccinated a high fraction of their population will be able to contain a Delta outbreak. By analyzing the fraction of newly vaccinated individuals and the number of COVID-19 deaths in 50 countries around the world, we observed a significant negative correlation between recent vaccination rate and COVID-19 deaths (R= -0.47, p-value = 0.0005) (Figure 2a). We further validated this result by performing a similar analysis on 427 counties across the United States of America (USA), and observed a similar negative correlation (R = -0.43, p-value<2.2e-16) (Figure 2b).

**FIG. 2:**
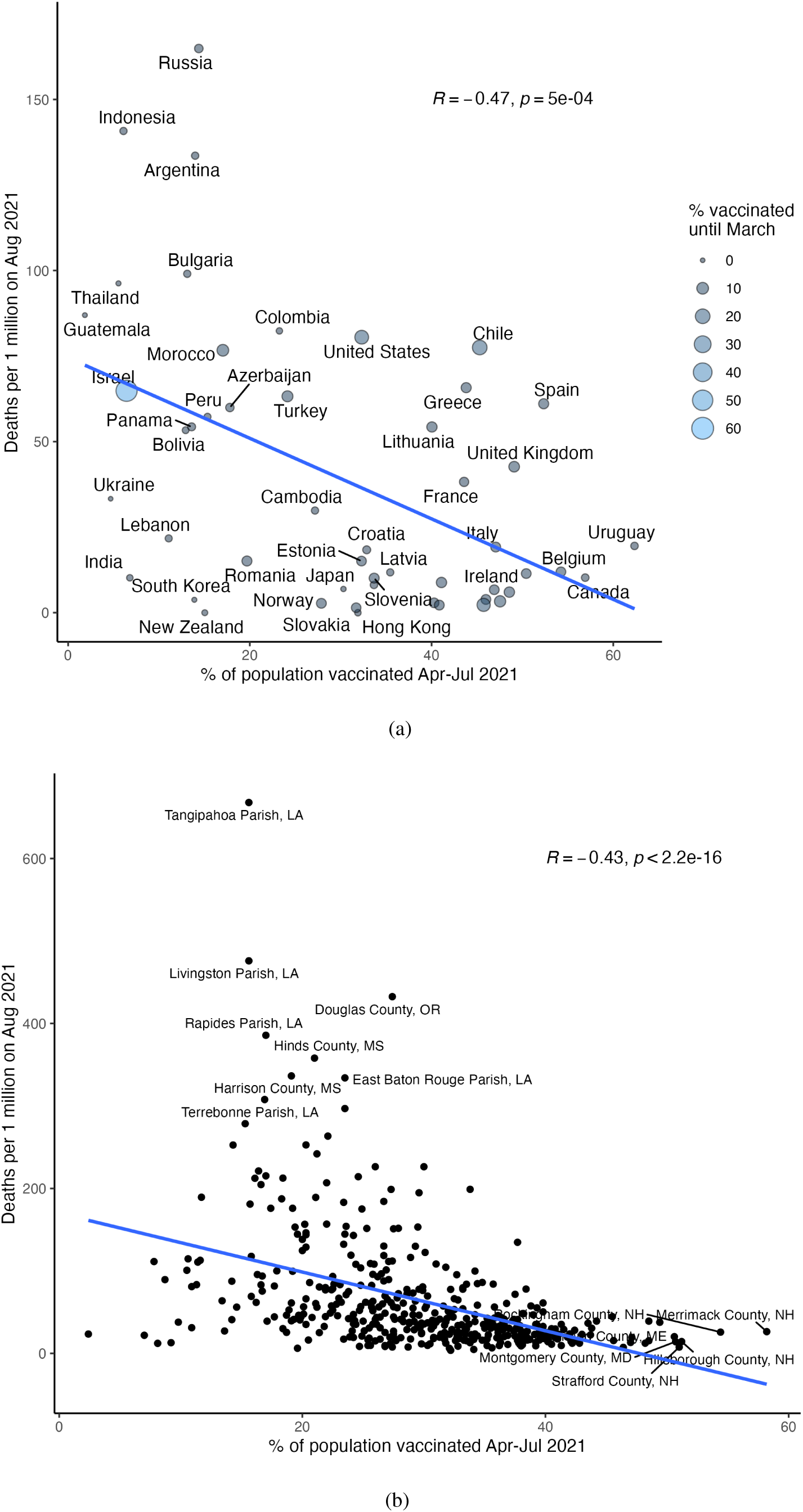
Effect of vaccinations on controlling Delta. The scatter plot shows the percent of population vaccinated in April to July 2021, which are expected to be effective in August vs. the number of deaths per 1 million individuals in each country (a) and USA county (b). Only countries with at population above 1 million people and counties with at least 100K people are presented). Size of dots is the percent of population vaccinated before April, expected to be less effective in August.

## DISCUSSION

In this study, we used a spatio-dynamic Monte Carlo algorithm to estimate the effect of the Delta variant on the COVID-19 outbreak that started in Israel in June 2021. We compared four different models with a combination of varying VE and theoretical reproduction rate. Based on the analysis, both the introduction of Delta and its higher reproduction rate as well as a time-decline in VE can explain this outbreak. Interestingly, we observe that the time-decline VE is responsible for the fact that the first to be infected are those vaccinated first, which are most prominently older populations, who are also at higher risk for severe disease. It is therefore recommended that counties that have vaccinated their elderly population in recent months monitor not just the number of confirmed cases, but also their age distribution and immunization status, and be aware of when this population will lose some of its immune defenses. Such monitoring will allow to detect in real-time the protective attenuation of the vaccine and provide this population another dose of the vaccine on time, which according to recent studies, has been found to be very effective against the delta variant [23, 24].

It is important to keep in mind that our models attempt to predict events that did not appear in reality. In the real world all the parameters of the model are inter-dependent. Increased reproduction rates might cause the government to impose restrictions sooner and cause a reduction in social interactions. A high reproduction rate may also result in more people getting vaccinated. As such, our analysis focuses on factors contributing to the June outbreak in Israel and its global implications, not on the prediction of cases in alternative scenarios.

Interestingly, when we examine the effect of *R*_*t*_ on the spread of the pandemic (Supplementary Figure 1) we find that in case of decline in the effectiveness of the vaccine against infection (which is the case today) the theoretical reproduction number, *R*_*t*_ should be relatively low (*R*_*t*_ < 2), in order not to push Israel below the HIT, which could be the case with a less contagious variant and/or presence of effective NPIs.

In summary, we argue that both the Delta variant higher infectiousness and the reduction of effectiveness of the vaccines contributed to pushing Israel below the HIT. If this is the case, the booster shots provided to all Israelis since August, are expected to reduce severe cases and the burden on hospitals, but not to bring Israel back to herd immunity without reinstating effective NPIs.

## Data Availability

All the data in available on:
1.https://datadashboard.health.gov.il/COVID-19/general
2. https://github.com/dancarmoz/israel_moh_covid_dashboard_data
3. https://ourworldindata.org/coronavirus

https://data.gov.il/dataset/covid-19

https://github.com/dancarmoz/israel_moh_covid_dashboard_data

## DATA AND CODE AVAILABILITY

All data used in this study is from public resources. The code to recreate figure 1 is available at https://github.com/hdeleon0/Delta-waning-.The code to recreate figure 2 is available at https://github.com/dviraran/covid_analyses/tree/master/delta.

## ACKNOWLEDGMENTS

DA is supported by the Azrieli Faculty Fellowship and is a Deloro Fellow. We thank Francesco Pederiva, Yair Lewis and Michael Geller for fruitful discussions.

## SUPPLEMENTARY FIGURES

**FIG. S.1:**
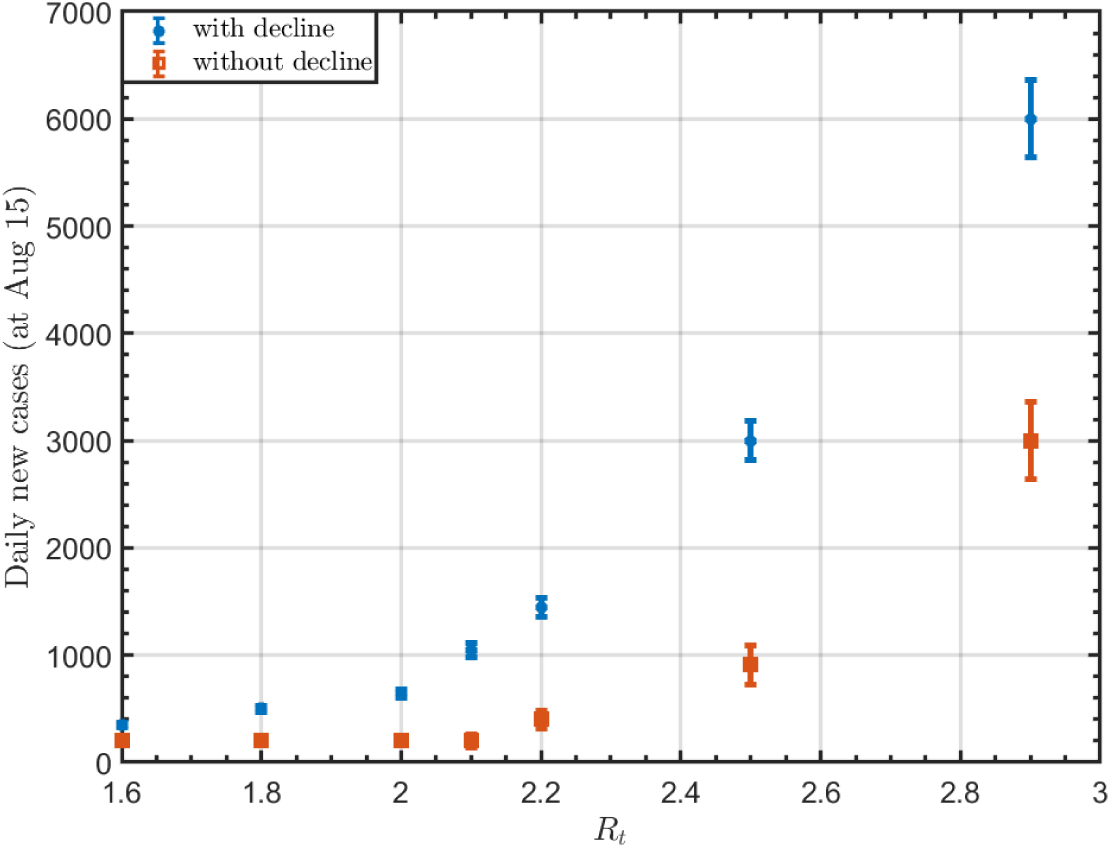
Predictions of total number of confirmed cases for August 15. Dots: The total number of confirmed cases in the presence of decline in the VE. Squares: The total number of confirmed cases without a decline in the VE. The error-bars are due to the uncertainty regarding the effectiveness of the vaccine and the numerical limitations of the calculation.

**FIG. S.2:**
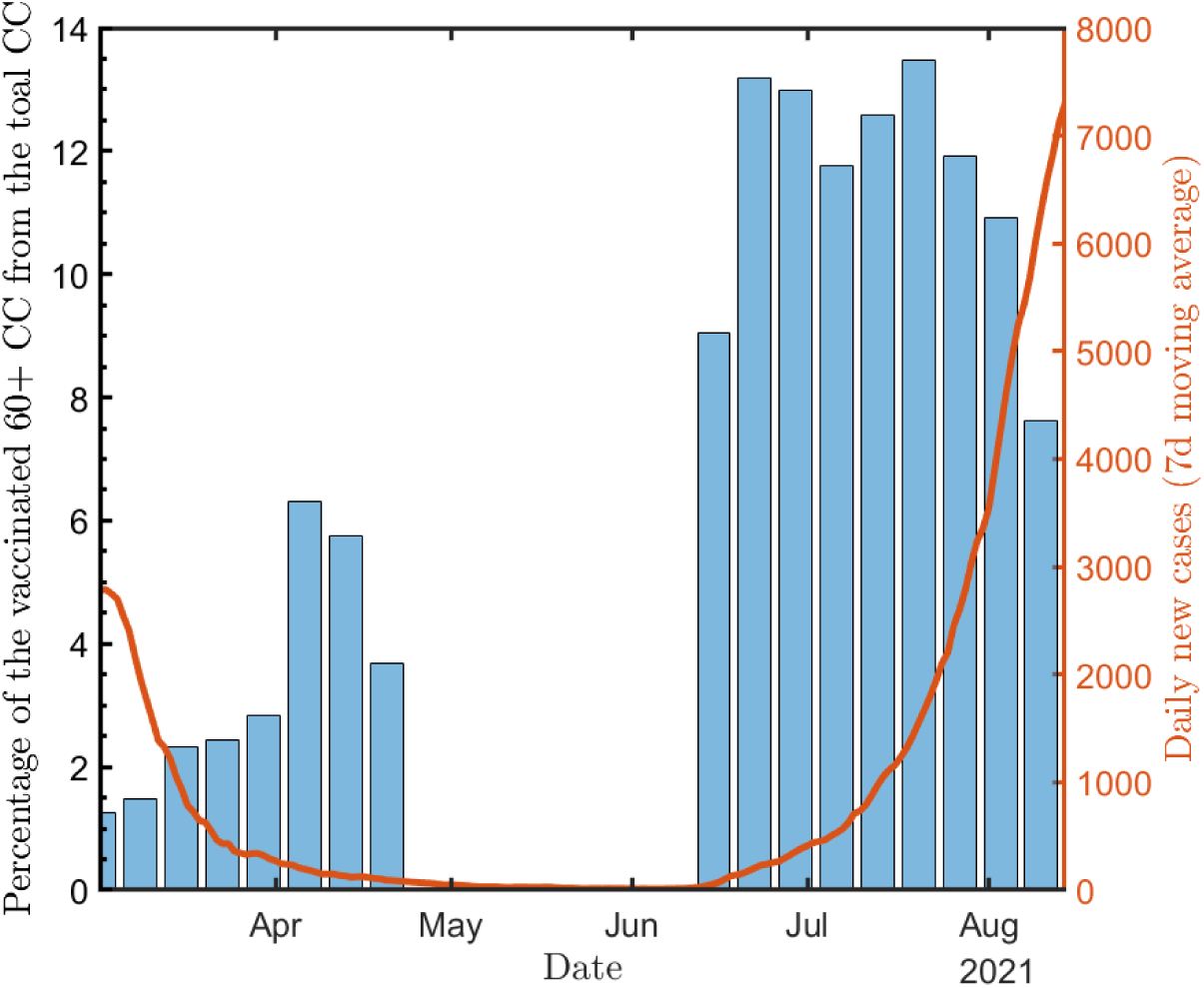
Bars: Percentage of verified vaccinated persons aged 60+ among all verified persons as of March 2021; Solid line: The daily total confirmed cases (all the data was taken from Refs. [5, 25]).

## References

[1] https://www.gisaid.org/hcov19-variants/.

[2] https://ourworldindata.org/.

[3] Ying Liu and Joacim Rocklöv. The reproductive number of the Delta variant of SARS-CoV-2 is far higher compared to the ancestral SARS-CoV-2 virus. Journal of Travel Medicine, 08 2021. taab124.

[4] Chun Huai Luo, C. Paul Morris, Jaiprasath Sachithanandham, Adannaya Amadi, David Gaston, Maggie Li, Nicholas J. Swanson, Matthew Schwartz, Eili Y. Klein, Andrew Pekosz, and Heba H. Mostafa. Infection with the sars-cov-2 delta variant is associated with higher infectious virus loads compared to the alpha variant in both unvaccinated and vaccinated individuals. medRxiv, 2021.

[5] https://data.gov.il/dataset/covid-19.

[6] Yair Goldberg, Micha Mandel, Yinon M. Bar-On, Omri Bodenheimer, Laurence Freedman, Eric J. Haas, Ron Milo, Sharon Alroy-Preis, Nachman Ash, and Amit Huppert. Waning immunity of the bnt162b2 vaccine: A nationwide study from israel. medRxiv, 2021.

[7] Ashley Fowlkes. Effectiveness of covid-19 vaccines in preventing sars-cov-2 infection among frontline workers before and during b. 1.617. 2 (delta) variant predominance—eight us locations, december 2020–august 2021. MMWR. Morbidity and Mortality Weekly Report, 70, 2021.

[8] Jennifer B Griffin. Sars-cov-2 infections and hospitalizations among persons aged 16 years, by vaccination status—los angeles county, california, may 1–july 25, 2021. MMWR. Morbidity and Mortality Weekly Report, 70, 2021.

[9] Hiam Chemaitelly, Patrick Tang, Mohammad R. Hasan, Sawsan AlMukdad, Hadi M. Yassine, Fatiha M. Benslimane, Hebah A. Al Khatib, Peter Coyle, Houssein H. Ayoub, Zaina Al Kanaani, Einas Al Kuwari, Andrew Jeremijenko, Anvar Hassan Kaleeckal, Ali Nizar Latif, Riyazuddin Mohammad Shaik, Hanan F. Abdul Rahim, Gheyath K. Nasrallah, Mohamed Ghaith Al Kuwari, Hamad Eid Al Romaihi, Adeel A. Butt, Mohamed H. Al-Thani, Abdullatif Al Khal, Roberto Bertollini, and Laith J. Abu-Raddad. Waning of bnt162b2 vaccine protection against sars-cov-2 infection in qatar. medRxiv, 2021.

[10] https://www.ncbi.nlm.nih.gov/pmc/articles/PMC3782273/.

[11] Hilla De-Leon and Francesco Pederiva. Particle modeling of the spreading of coronavirus disease (covid-19). Physics of Fluids, 32(8):087113, 2020.

[12] Hilla De-Leon and Francesco Pederiva. Statistical mechanics study of the introduction of a vaccine against covid-19 disease. Phys. Rev. E, 104:014132, Jul 2021.

[13] Omer Karin, Yinon M Bar-On, Tomer Milo, Itay Katzir, Avi Mayo, Yael Korem, Boaz Dudovich, Eran Yashiv, Amos J Zehavi, Nadav Davidovich, et al. Adaptive cyclic exit strategies from lockdown to suppress covid-19 and allow economic activity. MedRxiv, 2020.

[14] Nicholas G Davies, Adam J Kucharski, Rosalind M Eggo, Amy Gimma, W John Edmunds, Thibaut Jombart, Kathleen O’Reilly, Akira Endo, Joel Hellewell, Emily S Nightingale, et al. Effects of non-pharmaceutical interventions on covid-19 cases, deaths, and demand for hospital services in the uk: a modelling study. The Lancet Public Health, 5(7):e375–e385, 2020.

[15] Shilei Zhao and Hua Chen. Modeling the epidemic dynamics and control of covid-19 outbreak in china. Quantitative biology (Beijing, China), page 1, 2020.

[16] Barak Mizrahi, Roni Lotan, Nir Kalkstein, Asaf Peretz, Galit Perez, Amir Ben-Tov, Gabriel Chodick, Sivan Gazit, and Tal Patalon. Correlation of sars-cov-2 breakthrough infections to time-from-vaccine; preliminary study. medRxiv, 2021.

[17] Hilla De-Leon and Francesco Pederiva. Using a physical model and aggregate data from israel to estimate the current (july 2021) efficacy of the pfizer-biontech vaccine. medRxiv, 2021.

[18] Paul Elliott, David Haw, Haowei Wang, Oliver Eales, C Walters, K Ainslie, C Atchison, C Fronterre, P Diggle, A Page, et al. React-1 round 13 final report: exponential growth, high prevalence of sars-cov-2 and vaccine effectiveness associated with delta variant in england during may to july 2021. 2021.

[19] https://usafacts.org/visualizations/coronavirus-covid-19-spread-map/.

[20] https://data.cdc.gov/Vaccinations/COVID-19-Vaccinations-in-the-United-States-County/8xkx-amqh.

[21] Hagai Rossman, Smadar Shilo, Tomer Meir, Malka Gorfine, Uri Shalit, and Eran Segal. Covid-19 dynamics after a national immunization program in israel. Nature medicine, pages 1–7, 2021.

[22] H. De-Leon, R. Calderon-Margalit, F. Pederiva, Y. Ashkenazy, and D. Gazit. First indication of the effect of covid-19 vaccinations on the course of the outbreak in israel. medRxiv, 2021.

[23] Yinon M. Bar-On, Yair Goldberg, Micha Mandel, Omri Bodenheimer, Laurence Freedman, Nir Kalkstein, Barak Mizrahi, Sharon Alroy-Preis, Nachman Ash, Ron Milo, and Amit Huppert. Bnt162b2 vaccine booster dose protection: A nationwide study from israel. medRxiv, 2021.

[24] Matan Levine-Tiefenbrun, Idan Yelin, Hillel Alapi, Rachel Katz, Esma Herzel, Jacob Kuint, Gabriel Chodick, Sivan Gazit, Tal Patalon, and Roy Kishony. Viral loads of delta-variant sars-cov2 break-through infections following vaccination and booster with the bnt162b2 vaccine. medRxiv, 2021.

[25] https://github.com/dancarmoz/israel_moh_covid_dashboard_data.

